# Childhood immunisation coverage in rural and a tribal settings in southern India and assessing effect of the Covid19 pandemic

**DOI:** 10.64898/2026.07.24.26358371

**Authors:** Reshma Raju, Rohan Michael Ramesh, Selvakumar Prasad, Kulandaipalayam Natarajan Sindhu, Saravanakumar Puthupalayam Kaliappan, Kumudha Aruldas, Jayaprakash Muliyil, Mahesh Moorthy, Rajeev Zachariah Kompithra, Sitara S.R. Ajjampur

## Abstract

**Purpose:** Childhood immunisation witnessed reduced coverage and delays globally during the Covid19 pandemic. This study aimed to identify gaps in routine childhood immunisation in southern India and the effect of the pandemic in censused rural and tribal populations.

**Methods:** This study was conducted between August and September 2021 in a hard-to-reach tribal block (Jawadhu Hills) and rural (Timiri) block in Tamil Nadu. The proportion of full immunisation coverage (FIC), age-appropriate coverage and vaccination delays were calculated in the pre-pandemic (prior to March 2020) and pandemic (lockdown) period.

**Results:** Among 2746 children (aged <30 months, with immunisation cards available) surveyed in these populations, overall FIC among children between 12-23 months of age was 50.8% (95%CI: 46.6-55.0) in Jawadhu and 83.7%; (81.1-86.2) in Timiri. In Jawadhu, coverage of oral polio vaccine3 (OPV3) decreased from 78.3% (73.4-82.6) pre-pandemic to 70.0% (66.6-73.2) during the pandemic; inactivated polio vaccine2 (IPV2) from 76.9% (72.1-81.2) to 69.1% (65.5-72.4); Pentavalent3 from 78.0% (73.1-82.4) to 70.2% (66.8-73.5); and Measles containing vaccine1 (MCV1) from 74.6% (67.6-80.8) to 56.7% (53.3-60.1). In Timiri, only MCV1 coverage decreased from 92.9% (88.7-95.9) to 85.4% (83.3-87.3). Factors associated with not receiving MCV1 included due date of vaccination falling during the pandemic lock downs, higher birth order, younger and less educated parents, and residing in the tribal block.

**Conclusion:** This study highlighted differences in routine childhood immunisation vaccine coverage and impact of the pandemic in a geographically remote tribal block in contrast to a more accessible, rural block. Resilient health systems are essential to ensure sustainability in gains in vaccination coverage, especially in remote areas, and should be part of pandemic preparedness.

## INTRODUCTION

Immunisation is one of the most cost-effective public health interventions, preventing 4-5 million child deaths annually (1). Full Immunisation Coverage (FIC) (proportion of 12-23-month-olds receiving Bacille Calmette-Guérin (BCG), three polio doses, diphtheria, tetanus toxoid and pertussis (DTP3) vaccine and one measles dose) determines program performance (2). The “Immunisation Agenda 2030”, launched by the World Health Assembly in 2020, aims to reach 90% essential immunisation coverage, halve the number of unimmunised children (3). Global coverage declined during the COVID-19 pandemic, with nearly 23 million children were not fully immunised in 2020, an increase from 2019 (4). Globally, DPT3/polio coverage fell from 86% in 2019 to 83% in 2020, and MCV1 from 86% to 84% (5).

India’s Universal Immunisation Programme provides free vaccination against nine vaccine-preventable diseases (VPDs) nationwide (diphtheria, pertussis, tetanus, Hepatitis B, Haemophilus influenza type B (given as Pentavalent vaccine), poliomyelitis, measles, rubella and childhood tuberculosis), with Japanese encephalitis, rotavirus, and pneumococcal vaccines added sub-nationally. Mission Indradhanush and later Intensified Mission Indradhanush were launched to improve FIC to 90% in underserved and tribal areas (6), while Tamil Nadu’s Dr. Muthulakshmi Reddy Maternity Benefit Scheme (MRMBS) supports institutional delivery and child immunisation through maternal incentives through wage compensation in instalments (140 to 220 USD) (7).

Before the pandemic, immunisation in India was delivered typically through health sub-centres (HSC) and Integrated Child Development Services Centres (ICDS) by anganwadi workers (AWW), accredited social health activists (ASHA) and village health nurses (VHN) (8). FIC among children aged 12-23 months increased from 62% in NFHS-4 to 76% in NFHS-5, and from 69.7% to 89.2% in Tamil Nadu, but NFHS data do not separate pre-pandemic from pandemic periods (9).

During COVID-19, routine immunisation was disrupted by lockdowns, travel restrictions, staff shortages, and other service barriers (10,11). Paediatricians reported challenges including reduced immunisation supplies, protective equipment shortages, parents’ financial constraints, and low awareness of service availability (12). The pandemic’s effect on routine childhood vaccination in India remains poorly studied, with few community-based studies from rural areas (13). This study aimed to determine routine immunisation coverage and delays and assess the impact of the Covid-19 pandemic by conducting a door-to-door survey in a tribal and rural block in Tamil Nadu, India.

## MATERIALS AND METHODS

### Study setting

This study was conducted in Jawadhu Hills block of Thiruvannamalai district and Timiri block of Ranipet district in Tamil Nadu. Jawadhu Hills is a tribal block with indigenous communities (categorised as scheduled tribes by Government of India), comprising 154 villages, two PHCs, nine HSCs, and a population of 32,813, located on hilly terrain with poor connectivity in a reserve forest (14). Timiri block has 226 villages with four primary health centres (PHCs), 27 health subcentres (HSCs) and a population of 1,08,119.

### Study design and data collection

This cross-sectional survey was conducted between August-September 2021 with parents of children born between March 2019 and February 2021 using a WHO-adapted questionnaire administered by trained staff after informed consent (15) (**Supplementary file: Figure 1**). Children who were not alive or were stillborn were excluded. Data collected included birth details, immunisation status, and parental demographics. Immunisation records were obtained from immunisation cards, with missing data noted. Children with documented vaccination dates in their immunisation cards were considered vaccinated, while those without were considered unvaccinated. Reasons for delayed or missed immunisations were documented. Data was captured using SurveyCTO mobile software (Dobility, Inc., Cambridge, Massachusetts).

**Figure 1A and 1B:**
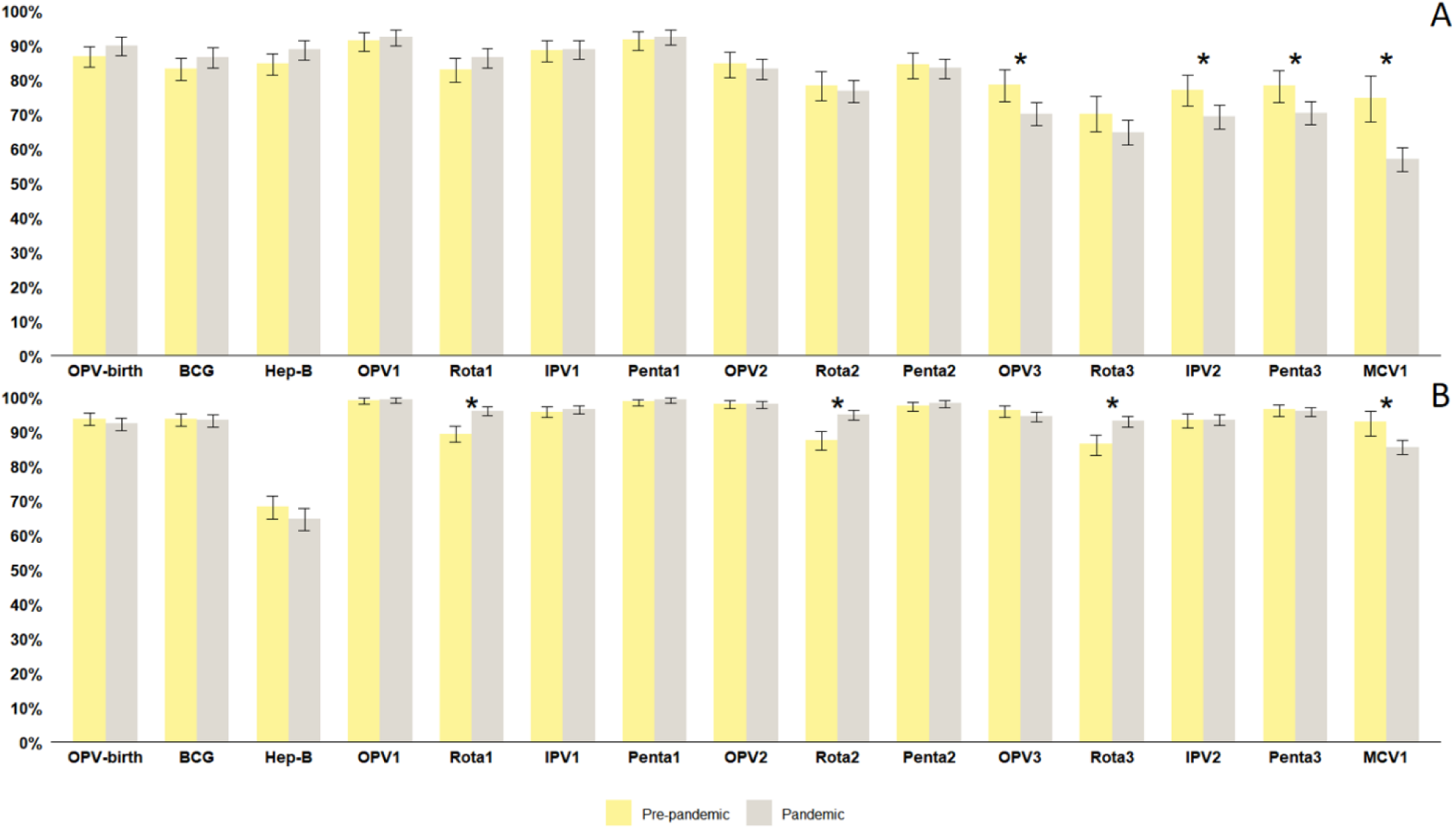
Individual vaccine coverage proportions in Jawadhu Hills (1A) and Timiri (1B) stratified by pre-pandemic and pandemic period (*indicates significant difference between the pandemic and the pre-pandemic period)

### Statistical analysis

Operational definitions are provided in **Supplementary Table 1**. Vaccines/doses were classified as pre-pandemic or pandemic if the expected vaccination date fell before or after 17 March 2020. Full immunisation coverage (FIC) was estimated among children aged 12-23 months, with stratified FIC for those whose eligible doses fell entirely before or after the cut-off. Descriptive statistics were expressed as proportions with 95% CIs, and differences between groups were tested using chi-square or Fisher’s exact tests. Age-appropriate coverage and non-receipt were estimated using unadjusted ORs with 95% CIs, delay proportions were similarly reported, and multivariable logistic regression identified factors associated with not receiving MCV1 and PV3 (aORs, 95% CIs); p<0.05 was considered significant. Analyses were performed in STATA 16.0 and figures created in R v4.1.0 (ggplot2).

### Ethical considerations

The Institutional Review Board (IRB) of the Christian Medical College, Vellore approved the study and written, informed consent was obtained in the local language from the parent, or the primary caregiver by trained field workers prior to administration of the survey.

## RESULTS

### Study population characteristics

Between March 2019 to February 2021, 4290 births (1563 in Jawadhu block and 2727 in Timiri block) were identified and among these, 3216 (75.0%) children (1254 in Jawadhu and 1962 in Timiri) participated in the study. Non-participation was due to temporary migration (6.1% in Jawadhu and 16.8% in Timiri), permanent migration (0.8% in Jawadhu and 6.9% in Timiri), doorlocks (10.9% in Jawadhu and 1.9% in Timiri), death (2% in Jawadhu and 2.2% in Timiri) and a few refusal to participate (none in Jawadhu and 0.1% in Timiri). Among the children who participated, 85.4% (2746/3216, 1083 in Jawadhu and 1663 in Timiri) had immunisation cards and were included in further analysis. Sociodemographic characteristics were similar in both groups (**Supplementary file: Table 2)**. Mothers were the main respondents (96.4%; mean age 25.6 years). Among children, 28.0% were <1 year, 51.0% were 12-24 months, and 21.0% were >24 months. Female children comprised 47.5% (1304). Government facility births were more common in Jawadhu (88.5%) than in Timiri (72.2%). More children in Timiri block were first or second in birth order (91.2%) than in Jawadhu (68.1%). Parental education (>secondary education) was lower in Jawadhu (mothers: 16.2%; fathers: 21.6%) than Timiri (mothers: 63.3%; fathers: 51.4%) (**Supplementary file: Table 3**).

### Immunisation coverage before and during the pandemic in Jawadhu and Timiri

Age-appropriate coverage, delay, and non-vaccination stratified for pandemic and pre-pandemic period in both Timiri and Jawadhu are presented in Supplementary Table 4. Overall FIC was lower in Jawadhu (50.8%; 95% CI: 46.6%-55.0%) than in Timiri (83.7%; 81.1%-86.2%). In Jawadhu, FIC decreased during the pandemic period (38.4%; 33.2%-43.8%) compared to pre-pandemic levels (59.1%; 36.4%-79.3%). While there was a slight decrease in Timiri during the pandemic period (78.7%; 74.4%-82.5%) from pre-pandemic period (85.3%; 68.9%-95.0%) this was not significant.

In Jawadhu, documentation was missing for BCG in 15.3% (n=166), however a BCG scar was present in 65.0% (n=108) of those children. No documentation of the birth doses of OPV and Hepatitis B were seen in 11.9% and 13.6% of children, respectively during the study period. About one-third of the children did not receive all three doses of OPV and PV, 38.0% did not receive all three doses of RV, 32.2% did not receive two doses of IPV and 40.1% did not receive MCV1 during the study period. Coverage declined significantly in the pandemic period in Jawadhu **(Figure 1A**) for several vaccine doses. These included OPV3 which decreased from 78.3% (73.4-82.6) to 70.0% (66.6-73.2) (OR 1.55; 1.14-2.1), IPV2 which decreased from 76.9% (72.1-81.2) to 69.1% (65.5-72.4) (OR 1.49; 1.11-2.00), PV3 which decreased from 78.0% (73.1-82.4) to 70.2% (66.8-73.5) (OR 1.50; 1.11-2.03) and MCV1 which showed a steep decrease from 74.6% (67.6-80.8) to 56.7% (53.3-60.1) (OR 2.24; 1.56-3.21).

In Timiri, there was no documented evidence of receiving the birth doses BCG in 6.6% (n=111) of the children but a BCG scar was present in 77.5% (n=86) of those children. Birth doses of OPV and hepatitis B vaccine were not documented in the vaccine cards of 7.1% and 33.8% of the children, respectively, during the study period. Approximately 6% did not receive all three doses of OPV and PV, 9% did not receive two doses of IPV and 13% each did not receive all three doses of RV and MCV1 during the study period. In Timiri, only MCV1 declined significantly from 92.9% (88.7-95.9) to 85.4% (83.3-87.3) during the pandemic (OR 2.23; 1.31-3.80) **(Figure 1B**). Coverage of three doses of RV (introduced in 2017 in Tamil Nadu) increased from 81.0% (77.5-84.3) pre-pandemic to 89.6% (87.7-91.3) during the pandemic in Timiri (OR 0.49; 0.37-0.66), with no significant differences for the other vaccines.

### Factors associated with not receiving MCV1 and Pentavalent Vaccine 3

Multivariable logistic regression was performed to determine the factors associated with low coverage of MCV1 and PV3. Not receiving MCV1 was associated with: i) due date of vaccination during pandemic period (aOR 2.33; 95% CI:1.71-3.18), ii) birth order > 2 (aOR 1.44; 1.11-1.85), iii) having a younger mother (<20 years of age: aOR 1.68; 1.11-2.54), iv) having parents educated only up to the primary school level or below (mothers education: aOR 2.19; 1.68-2.83 and fathers education: aOR 1.66; 1.29-2.14) and v) residing in Jawadhu (aOR 2.35; 1.82-3.04) **(Figure 2A**). Factors associated with not receiving PV3 were similar (**Figure 2B**).

**Figure 2A and 2B:**
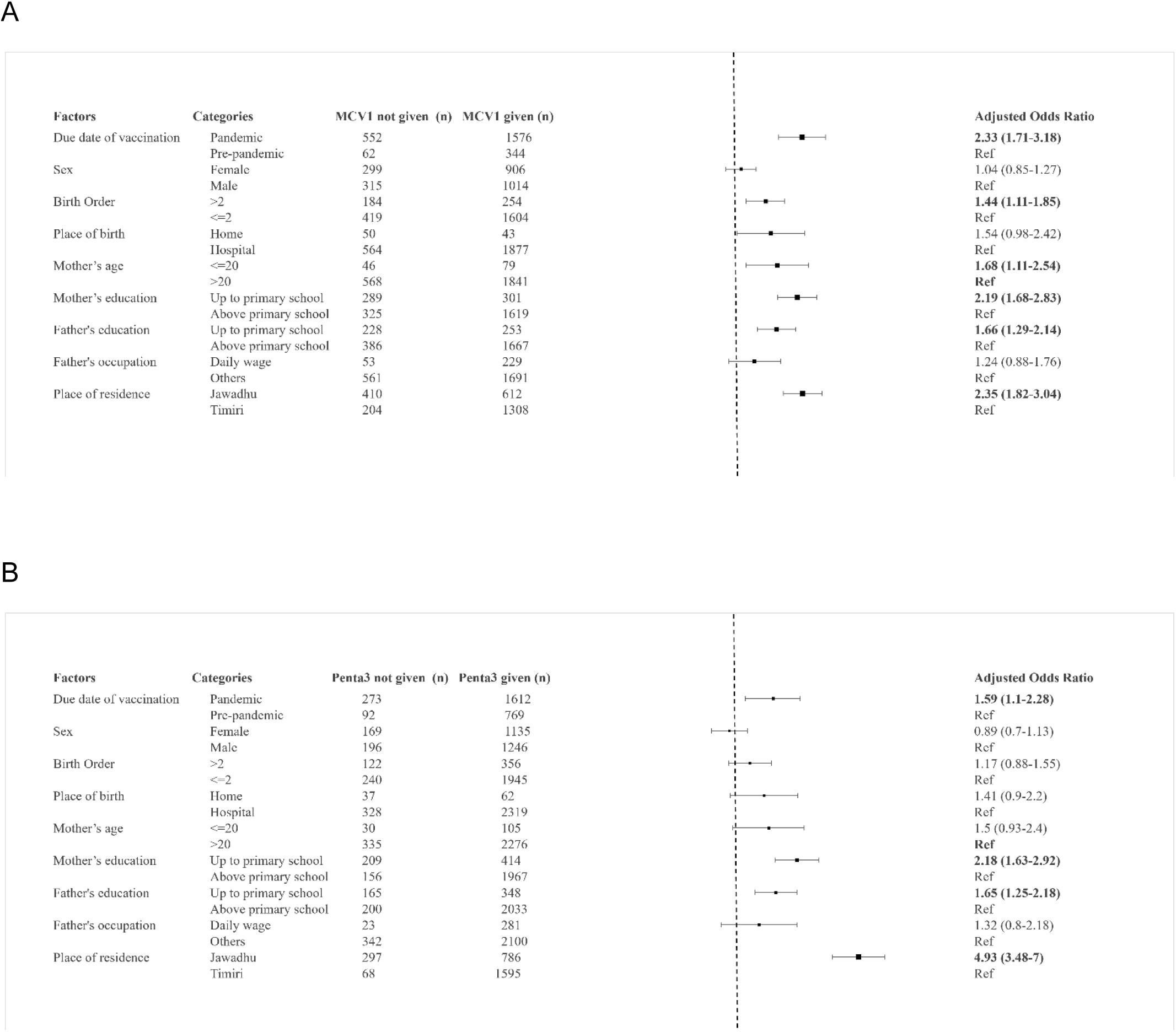
Logistic regression model to determine the risk factors for not receiving the first dose of measles containing vaccine (MCV1) (2A) and the third dose of Pentavalent (2B)

### Delayed in immunisation

In Jawadhu, among those who were vaccinated (n=1083), approximately 20-25% of all the doses between 6 weeks and 14 weeks (OPV, RV and PV), and 85% of the doses at 9 months, MCV1 were delayed during the study period **(Supplementary file: Table 5).** Delay in receiving the Hepatitis B birth dose occurred significantly more often during the pandemic (14.7%; 11.6-18.2) than in the pre-pandemic period (9.7%; 7.1-12.7) (OR 1.60; 1.06-2.45). During the pre-pandemic period, 30.8% (25.3-36.8) of the children received their IPV2 later than the recommended time and the proportion experiencing delay increased during the pandemic (40.9%; 36.6-45.3) (OR 1.55; 1.13-2.12) **(Figure 3A).** There were fewer delayed vaccinations in Timiri. Approximately 5-7% of all vaccine doses between 6 weeks and 14 weeks (OPV, RV and PV) were delayed but at 9 months, 61% of MCV1 was delayed **(Supplementary file: Table 5)**. Delays in OPV1 vaccination occurred less often during the pandemic (4.6%) compared to the pre-pandemic period (7.1%) (OR 0.64; 0.42-0.9). A similar trend was seen for IPV1, RV2 and PV2. The proportion of children who with delayed MCV1 was much higher than the other vaccines in Timiri, and here too it was higher in pre-pandemic (75.6%; 69.2-81.3) than pandemic period (59.0%; 56.0-61.9) (OR 0.46; 0.33-0.65) **(Figure 3B)**.

**Figure 3A and 3B:**
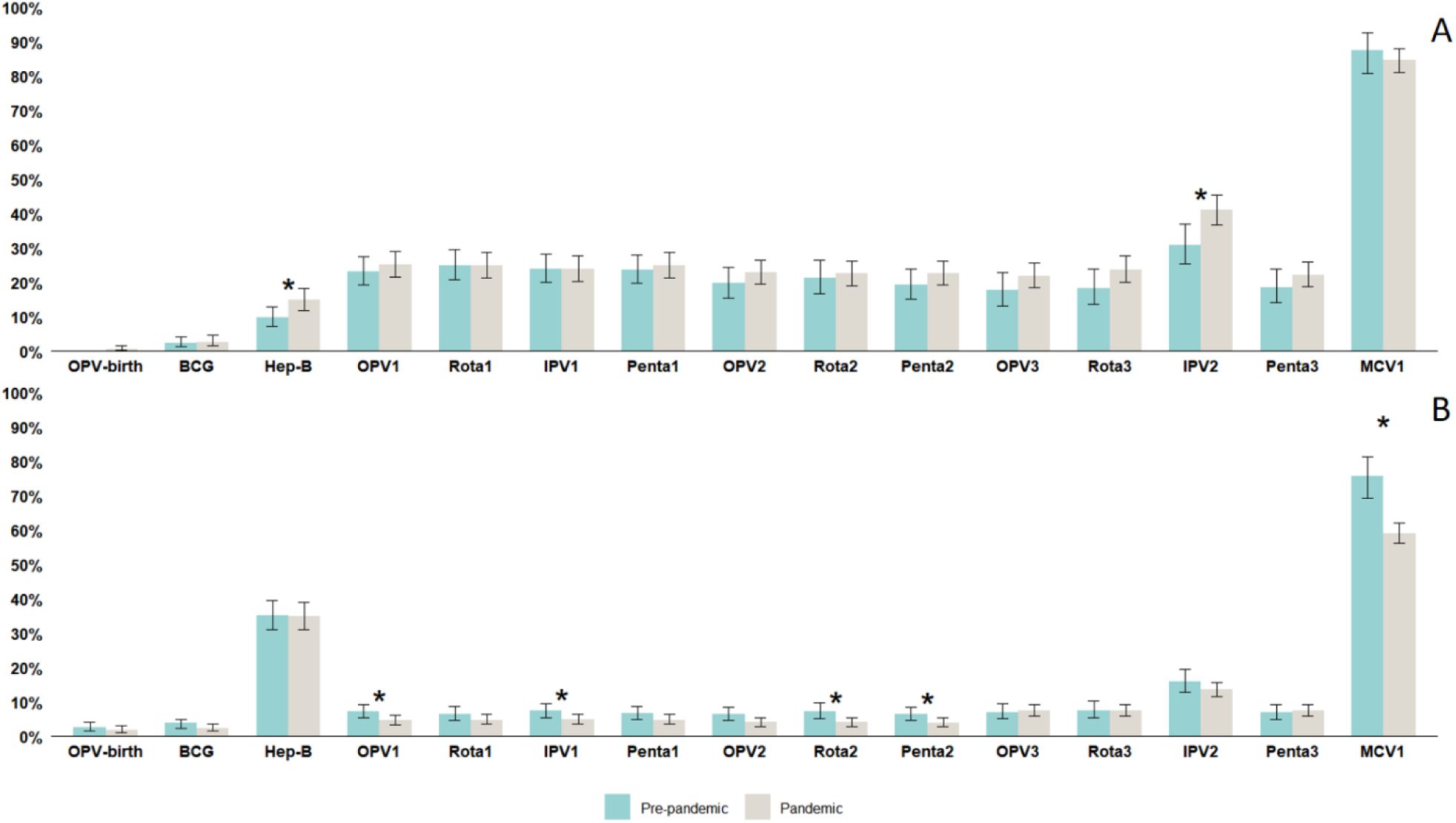
Proportion of delayed vaccination for individual vaccines in Jawadhu Hills (3A) and Timiri (3B) stratified by pre-pandemic and pandemic period (*indicates significant difference between the pandemic and the pre-pandemic period)

The most common reasons reported for missing the MCV1 vaccine were mother’s lack of awareness (30.7%), mother or child having an illness on the scheduled date of immunisation (22.2%), being out-of-station on the date of immunisation (13.8%), having a fear of Covid-19 (11.1%), poor accessibility (4%) and vaccine safety concerns (3.6%). The reasons for delaying MCV1 vaccine were also similar (**Supplementary file: Figures 2A and 2B**).

## DISCUSSION

During COVID-19, routine childhood immunisation declined in 73 countries, and 24.4 million children missed vaccinations in 2021, exceeding pre-pandemic levels by 6 million (19). Indian studies also showed reduced coverage: children born after COVID-19 had a lower chance of being vaccinated, and district-level analyses found declines in BCG, PV3, OPV3, and MCV1 coverage (20,21). These reductions likely reflected overburdened health systems and lockdowns (19). However, regional averages can hide major inequities in underserved communities. This study identified such immunisation coverage inequity in a tribal area located in a geographically and socioeconomically vulnerable area in southern India.

Vaccine coverage for doses beyond three months (OPV3, IPV2, Pentavalent3, and MCV1) was lower in the hard-to-reach tribal block during the pandemic than for doses given near birth. This suggests that pandemic effects may be greater in socioeconomically disadvantaged communities. A Brazilian study also found a 20% decline in childhood vaccinations during peak social distancing months, with poorer and underdeveloped regions most affected (22). In our tribal area, immunisation at birth and soon after was largely unaffected, likely because institutional deliveries were high (91%) with vaccines and staff were available. Parents may have prioritised early doses over later ones, and lower maternal education and lack of awareness were key reasons for missed later doses. Similarly, Nandi et al. found that delayed vaccination increased during the pandemic for DPT and OPV compared with COVID-unaffected children (20).

In the tribal hard-to-reach area, delayed vaccination was more common both before and during the pandemic than in the rural site. This suggests that national and regional coverage averages can mask inequities in disadvantaged communities unless they are closely monitored. It is important to note, cost of immunisation campaigns in hard-to-reach communities would be lower than managing VPD outbreaks (23). In India, where 50% of health expenditure is out-of-pocket (24), severe infections from missed vaccines would perpetuate poverty in these disadvantaged households and communities (23).

Measles is highly infectious, and missed MCV1 doses can trigger outbreaks. Global MCV1 coverage fell from 86% in 2019 to 81% in 2021, then rose to 83% in 2022 (19). Some Indian regions with low coverage reported measles outbreaks in 2022 (25). In this study, low MCV1 coverage was associated with vaccination due dates during the pandemic, higher birth order, maternal age under 20 years, primary-level parental education, and tribal residence. Bettampadi et al. found that maternal literacy improved FIC in India, with higher maternal education increasing the chance of full immunisation by 40% (26). A systematic review from Africa also found that maternal and paternal education, residence, and delivery location influenced immunisation coverage (27). Lower female literacy may indirectly contribute to higher birth order and younger maternal age, both linked to poor coverage in this study. Lockdowns, vaccine shortages, and poor caregiver recall across the five-month gap before MCV1 likely also reduced coverage. Phone reminders before due dates may help, as shown in southern India and Africa (28,29). In hard-to-reach areas, home visits by healthcare workers may be needed for unimmunised children.

The study had a few limitations. We did not perform an a priori sample size calculation and instead included all eligible children from the area. However, the achieved sample had >80% power to detect coverage differences of >=7% between the pre-pandemic and pandemic periods. The pre-pandemic FIC analysis among 12-23-month-old children included fewer observations (n=56), which may limit the robustness of block-level estimates. We excluded children without immunisation cards (15%) to reduce recall bias; although sociodemographic characteristics were similar, however some associated factors may still have been missed.

This study shows that remote areas are more vulnerable to disruptions in childhood immunisation, even in states with otherwise robust healthcare systems. Pandemic preparedness should therefore identify and plan for remote at-risk areas in more granular detail. Low parental education (especially maternal), was associated with lower childhood vaccination and should be addressed through policies that improve literacy, particularly female literacy, which benefits child survival and health in multiple ways. Telephone or text reminders before vaccination due dates may also improve uptake, especially for vaccines due at three months of age or later.

In conclusion, events such as pandemics, civil conflicts and natural disasters have the potential to rapidly reverse gains in vaccination coverage achieved over the years, and put millions of children at risk and health system preparedness and resilience planning should, as a priority, ensure childhood immunisation activities.

## Data Availability

The datasets generated and analyzed during the current study are available from the corresponding author on reasonable request

## Acknowledgement

We thank all the study participants and the communities who have participated or supported this study. We thank the field managers, Rajeshkumar Rajendiran and Chinnaduraipandi Paulsamy, and field supervisors and field workers, for providing logistic support for conducting the snakebite surveys and Naveenkumar for providing support with data capture forms. We acknowledge the work of the members of the DeWorm3 study teams and affiliated institutions.

## Author contributions

Conceptualization: SSRA, RMR, KA, KNS, SPK, RZK; Methodology: RMR, KA, SPK, RZK; Data collection: RMR, SPK, RR; Formal analysis: RR, SP, MM, RMR, JPM; Validation: RR, SP, RMR, SPK; Visualization: SP, RR, MM; Manuscript writing: RR, SSRA; Reviewing and editing: SSRA, RMR, RR, KA, MM, KNS, SPK, RZK, JPM.

## Declaration of AI in scientific writing

During preparation of the draft manuscript, the authors used Paperpal software (Editage) to reduce word count. After using this software, the author(s) reviewed and edited the final draft as needed and take full responsibility for the content of the publication.

## Ethical Approval

The Institutional Review Board (IRB) of the Christian Medical College, Vellore approved the study.

## Supplementary Tables Legends

Supplementary Table 1: Operational defintions

Supplementary Table 2: Basic sociodemographic profile of participants with and without an immunisation card

Supplementary Table 3: Socio-demographic profile of the study participants in Jawadhu Hills and Timiri

Supplementary Table 4: Proportion of children with age-appropriate vaccination delayed vaccination and who are unvaccinated stratified for pandemic and per-pandemic period.

Supplementary Table 5: Overall delay in vaccination at 6^th^,10^th^ and 14^th^ week doses in Jawadhu Hills and Timiri during the study period

## Supplementary Figures Legends

Supplementary Figure 1: Flowchart of the participant enrolment process

Supplementary Figure 2A and 2B: Reasons for missed MCV1 dose (A) and delayed MCV1 dose (B) in Jawadhu Hills and Timiri together.

