## Supplement tables and figures for "Childhood immunisation coverage in rural and a tribal settings in southern India and assessing effect of the Covid19 pandemic"

**Vaccine coverage - Supplementary Tables**

**Supplementary table 1: Operational definitions**

| Variable / definition | Operational definition |
|---|---|
| Expected date of vaccination | Calculated from the child's date of birth and the routine immunisation schedule. |
| Age at vaccination | Actual vaccination date minus date of birth. |
| Age-appropriate vaccination coverage | proportion of vaccinated children among those eligible. |
| Immunisation delay | Difference between expected and actual vaccination dates. |
| Pandemic classification | Vaccines/doses were classified as pre-pandemic or pandemic based on whether the expected vaccination date fell before or after 17 March 2020. |
| Full immunisation coverage (FIC) | Calculated among children aged 12–23 months. |
| Stratified FIC | Children expected to receive all their eligible vaccine (BCG, three doses of OPV, three doses of PV, and MCV1) before 17$^{th}$ March 2020 contributed to the pre-pandemic FIC calculation, and those expected to receive all their eligible vaccines after 17$^{th}$ March 2020 contributed to the pandemic FIC calculation |
| Hepatitis B delay | Birth dose given after 24 hours. |
| BCG / zero-dose OPV delay | Dose given after 28 days of birth. |
| Pentavalent / OPV / RV delay | First dose given after 70 days; second and third doses more than 28 days after the previous dose. |
| MCV1 delay | Dose given after 280 days of birth. |

**Supplementary Table 2**: Basic sociodemographic profile of participants with and without an immunization card

| | Category | Immunization card available | Immunization card not available | p value |
|---|---|---|---|---|
| | | n (%) | n (%) | |
| **Age of the children** | <=1 year (n=885) | 750 (28.2) | 135 (24.4) | 0.072 |
| | >1 year (n=2331) | 1913 (71.8) | 418 (75.6) | |
| **Sex** | Female (n=1539) | 1268 (47.6) | 271 (49.0) | 0.552 |
| | Male (n=1677) | 1395 (52.4) | 282 (51.0) | |
| **Mother's age** | <=20 (n=166) | 134 (5.0) | 32 (5.8) | 0.465 |
| | >20 (n=3050) | 2529 (95.0) | 521 (94.2) | |
| **Mother's education** | Up to primary (n=733) | 622 (23.4) | 111 (20.1) | 0.094 |
| | Above primary (n=2483) | 2041 (76.4) | 442 (79.9) | |
| **Father's education** | Up to primary (n=610) | 512 (19.2) | 98 (17.7) | 0.411 |
| | Above primary (n=2606) | 2151 (80.8) | 455 (82.3) | |

**Supplementary Table 3**: Socio-demographic profile of the study participants in Jawadhu and Timiri

| Variables | Category | Jawadhu Hills | Timiri | Total |
|---|---|---|---|---|
| | | n (%) | n (%) | n (%) |
| **Age categories** (Timiri-1663; Jawadhu-1083) | <= 1 year | 271 (25·0) | 499 (30·0) | 770 (28·0) |
| | > 1 to <= 2 year | 563 (52·0) | 836 (50·3) | 1399 (51·0) |
| | >2 years | 249 (23·0) | 328 (19·7) | 577 (21·0) |
| **Sex** (n=2476) Timiri-1663; Jawadhu-1083 | Male | 550 (50·8) | 892 (53·6) | 1442 (52·5) |
| | Female | 533 (49·2) | 771 (46·4) | 1304 (47·5) |
| **Birth order** (n= 2663) (Timiri-1580; Jawadhu-1083) | <=2 | 737 (68·1) | 1448 (91·7) | 2185 (82·1) |
| | >2 | 346 (32·0) | 132 (8·4) | 478 (18·0) |
| **Place of birth** (n=2740) (Timiri-1657; Jawadhu-1083) | Private facility | 27 (2·5) | 459 (27·7) | 486 (17·7) |
| | Government facility | 958 (88·5) | 1197 (72·2) | 2155 (78·7) |
| | Home | 98 (9·1) | 1 (0·1) | 99 (3·6) |
| **Mother's age** (in years) (n=2746) (Timiri-1663; Jawadhu-1083) | <=20 | 81 (7·5) | 54 (3·3) | 135 (4·9) |
| | >20 | 1002 (92·5) | 1609 (96·8) | 2611 (95·1) |
| **Mother's education** (n=2746) (Timiri-1663; Jawadhu-1083) | Up to primary school | 553 (51·1) | 70 (4·2) | 623 (22·7) |
| | Middle school | 216 (19·9) | 134 (8·1) | 350 (12·7) |
| | Secondary school | 139 (12·8) | 407 (24·5) | 546 (19·9) |

| | | | | |
|---|---|---|---|---|
| | Above secondary | 175 (16·2) | 1052 (63·3) | 1227 (44·7) |
| **Father's education** (n=2746) (Timiri-1663; Jawadhu-1083) | Up to primary school | 423 (39·1) | 90 (5·4) | 513 (18·7) |
| | Middle school | 241 (22·3) | 253 (15·2) | 494 (18·0) |
| | Secondary school | 185 (17·1) | 465 (28·0) | 650 (23·7) |
| | Above secondary | 234 (21·6) | 855 (51·4) | 1089 (39·7) |
| **Father's occupation** (n=2746) (Timiri-1663; Jawadhu-1083) | Daily wages | 20 (1·9) | 284 (17·1) | 304 (11·1) |
| | Landowner | 873 (80·6) | 107 (6·4) | 980 (35·7) |
| | Business | 38 (3·5) | 106 (6·4) | 144 (5·2) |
| | Skilled | 111 (10·3) | 1004 (60·4) | 1115 (40·6) |
| | Government | 30 (2·8) | 106 (6·4) | 136 (5·0) |
| | Professional | 11 (1·0) | 37 (2·2) | 48 (1·8) |
| | Others | 0 (0) | 19 (1·1) | 19 (0·7) |

**Supplementary Table 4**: Proportion of children with age-appropriate vaccination delayed vaccination and who are unvaccinated stratified for pandemic and per-pandemic period.

| Age | Vaccine | Category | TIMIRI | | | | | | JAWADHU | | | | | |
|---|---|---|---|---|---|---|---|---|---|---|---|---|---|---|
| | | | Age appropriate vaccination | | Delayed vaccinated | | Unvaccinated | | Age appropriate vaccination | | Delayed vaccinated | | Unvaccinated | |
| | | | n | % (95% CI) | n | % (95% CI) | n | % (95% CI) | n | % (95% CI) | n | % (95% CI) | n | % (95% CI) |
| Birth | OPV birth dose | Pandemic (Timiri n=893; Jawadhu n=531) | 809 | 90·6 (88·5 - 92·4) | 15 | 1·7 (0·9 - 2·8) | 69 | 7·7 (6·1 - 9·7) | 474 | 89·3 (86·3 - 91·8) | 2 | 0·4 (0 - 1·4) | 55 | 10·4 (7·9 - 13·3) |
| | | Pre-Pandemic (Timiri n=770; Jawadhu n=552) | 702 | 91·2 (88·9 - 93·1) | 19 | 2·5 (1·5 - 3·9) | 49 | 6·4 (4·7 - 8·3) | 478 | 86·6 (83·5 - 89·3) | 0 | 0 (0) | 74 | 13·4 (10·7 - 16·5) |
| | BCG | Pandemic (Timiri n=893; Jawadhu **n=531**) | 813 | 91·0 (89·0 - 92·8) | 19 | 2·1 (1·3 - 3·3) | 61 | 6·8 (5·3 - 8·7) | 447 | 84·2 (80·8 - 87·2) | 12 | 2·3 (1·2 - 3·9) | 72 | 13·6 (0·8 - 16·8) |
| | | Pre-Pandemic (Timiri n=770 ; Jawadhu **n=552**) | 693 | 90·0 (87·7 – 92·0) | 27 | 3·5 (2·3 - 5·1) | 50 | 6·5 (4·9 - 8·5) | 448 | 81·2 (77·6 - 84·3) | 10 | 1·8 (0·9 - 3·3) | 94 | 17·0 (14·0 - 20·4) |
| | Hep B birth dose | Pandemic (Timiri n=893 ; Jawadhu **n=531**) | 375 | 42·0 (38·7 - 45·3) | 201 | 22·5 (19·8 - 25·4) | 317 | 35·5 (32·4 - 38·7) | 401 | 75·5 (71·6 - 79·1) | 69 | 13·0 (10·3 - 16·2) | 61 | 11·5 (8·9 - 14·5) |
| | | Pre-Pandemic (Timiri n=770; Jawadhu **n=552**) | 340 | 44·3 (40·6 - 47·7) | 184 | 23·9 (20·9 - 27·1) | 246 | 32·0 (28·7 - 35·4) | 421 | 76·3 (72·5 - 79·8) | 45 | 8·2 (6 - 10·8) | 86 | 15·6 (12·7 - 18·9) |
| 6weeks | OPV1 | Pandemic (Timiri n=961; Jawadhu **n=612**) | 908 | 94·5 (92·8 - 95·8) | 44 | 4·6 (3·3 - 6·1) | 9 | 0·9 (0·4 - 1·8 | 423 | 69·1 (65·3 - 72·8) | 141 | 23·0 (19·8 - 26·6) | 48 | 7·8 (5·8 - 10·3) |
| | | Pre-Pandemic (Timiri n=702; Jawadhu **n=471**) | 646 | 92·0 (89·8 - 93·9) | 49 | 7·0 (5·2 - 9·1) | 7 | 1·0 (0·4 – 2·0) | 330 | 70·1 (65·7 - 74·2) | 99 | 21·0 (17·4 – 25·0) | 42 | 8·9 (6·5 - 11·9) |

| | | | | | | | | | | | | | | |
|---|---|---|---|---|---|---|---|---|---|---|---|---|---|---|
| | **Rota1** | Pandemic (Timiri n=961; Jawadhu **n=612**) | 879 | 91·5 (89·5 - 93·2) | 43 | 4·5 (3·3 – 6·0) | 39 | 4·1 (2·9 - 5·5) | 397 | 64·9 (60·9 - 68·7) | 131 | 21·4 (18·2 - 24·9) | 84 | 13·7 (11·1 - 16·7) |
| | | Pre-Pandemic (Timiri n=702; Jawadhu **n=471**) | 586 | 83·5 (80·5 - 86·1) | 41 | 5·8 (4·2 - 7·8) | 75 | 10·7 (8·5 - 13·2) | 293 | 62·2 (57·7 - 66·6) | 97 | 20·6 (17 - 24·5) | 81 | 17·2 (13·9 - 20·9) |
| | **IPV1** | Pandemic (Timiri n=961 Jawadhu **n=612**) | 882 | 91·8 (89·9 - 93·4) | 44 | 4·6 (3·3 - 6·1) | 35 | 3·6 (2·5 - 5·0) | 414 | 67·7 (63·8 - 71·3) | 129 | 21·1 (17·9 - 24·5) | 69 | 11·3 (8·9 - 14·1) |
| | | Pre-Pandemic (Timiri n=702; Jawadhu **n=471)**) | 623 | 88·8 (86·2 – 91·0) | 49 | 7·0 (5·2 - 9·1) | 30 | 4·3 (2·9 - 6·0) | 317 | 67·3 (62·9 - 71·5) | 99 | 21·1 (17·4 - 25·0) | 55 | 11·7 (8·9 - 14·9) |
| | **Penta1** | Pandemic (Timiri n=961; Jawadhu **n=612)** | 907 | 94·4 (92·7 - 95·8) | 45 | 4·7 (3·4 - 6·2) | 9 | 0·9 (0·4 - 1·8) | 425 | 69·4 (65·6 - 73·1) | 140 | 22·9 (19·6 - 26·4) | 47 | 7·7 (5·7 - 10·1) |
| | | Pre-Pandemic (Timiri n=702; Jawadhu **n=471)** | 646 | 92·0 (89·8 - 93·9) | 46 | 6·6 (4·8 - 8·6) | 10 | 1·4 (0·7 - 2·6) | 329 | 69·9 (65·5 – 74) | 101 | 21·4 (17·8 - 25·4) | 41 | 8·7 (6·3 - 11·6) |
| **10 weeks** | **OPV2** | Pandemic (Timiri n=1026; Jawadhu **n=690)** | 962 | 93·8 (92·1 - 95·2) | 41 | 4 (2·9 - 5·4) | 23 | 2·2 (1·4 - 3·3) | 443 | 64·2 (60·5 - 67·8) | 130 | 18·8 (16·0 – 22·0) | 117 | 17·0 (14·2 – 20·0) |
| | | Pre-Pandemic (Timiri n=637; Jawadhu **n=393)** | 585 | 91·8 (89·4 - 93·8) | 39 | 6·1 (4·4 - 8·3 | 13 | 2·0 (1·1 - 3·5) | 267 | 67·9 (63·1 - 72·5) | 65 | 16·5 (13·0 - 20·6) | 61 | 15·5 (12·1 - 19·5) |
| | **Rota2** | Pandemic (Timiri n=1026; Jawadhu **n=690)** | 935 | 91·1 (89·2 - 92·8) | 39 | 3·8 (2·7 - 5·2) | 52 | 5·1 (3·8 - 6·6) | 410 | 59·4 (55·7 - 63·1) | 118 | 17·1 (14·4 - 20·1) | 162 | 23·5 (20·4 - 26·8) |
| | | Pre-Pandemic (Timiri n=637)**;** Jawadhu **n=393)** | 517 | 81·2 (77·9 - 84·1) | 40 | 6·3 (4·5 - 8·5) | 80 | 12·6 (10·1 - 15·4) | 242 | 61·6 (56·6 - 66·4) | 65 | 16·5 (13·0 - 20·6) | 86 | 21·9 (17·9 - 26·3) |
| | **Penta2** | Pandemic (Timiri n=1025) ; Jawadhu **n=691)** | 966 | 94·2 (92·6 - 95·6 | 39 | 3·8 (2·7 - 5·2) | 20 | 2·0 (1·2 - 3·0) | 445 | 64·4 (60·7 – 68·0) | 129 | 18·7 (15·8 - 21·8) | 117 | 16·9 (14·2 - 19·9) |
| | | Pre-Pandemic (Timiri n=638)**;** Jawadhu **n=392)** | 582 | 91·2 (88·8 - 93·3 | 39 | 6·1 (4·4 - 8·3) | 17 | 2·7 (1·6 - 4·2) | 267 | 68·1 (63·2 - 72·7) | 63 | 16·1 (12·6 - 20·1) | 62 | 15·8 (12·3 - 19·8) |

| | | | | | | | | | | | | | | |
|---|---|---|---|---|---|---|---|---|---|---|---|---|---|---|
| **14 weeks** | **OPV3** | Pandemic (Timiri n=1130)**;** Jawadhu **n=756)** | 987 | 87·4 (85·3 - 89·2) | 80 | 7·1 (5·7 - 8·7) | 63 | 5·6 (4·3 - 7·1) | 414 | 54·8 (51·1 - 58·4) | 115 | 15·2 (12·7 - 18) | 227 | 30·0 (26·8 - 33·4) |
| | | Pre-Pandemic (Timiri n=533; Jawadhu **n=327)** | 476 | 89·3 (86·4 - 91·8) | 36 | 6·8 (4·8 - 9·2) | 21 | 3·9 (2·5 - 6·0) | 211 | 64·5 (59·1 - 69·7) | 45 | 13·8 (10·2 - 18) | 71 | 21·7 (17·4 - 26·6) |
| | **Rota3** | Pandemic (Timiri n=1125; Jawadhu **n=753)** | 968 | 86·0 (83·9 – 88·0) | 78 | 6·9 (5·5 - 8·6) | 79 | 7·0 (5·6 - 8·7) | 372 | 49·4 (45·8 – 53) | 114 | 15·1 (12·7 - 17·9) | 267 | 35·5 (32·0 - 39·0) |
| | | Pre-Pandemic (Timiri n=538; Jawadhu **n=330)** | 429 | 79·7 (76·1 - 83·1) | 35 | 6·5 (4·6 - 8·9) | 74 | 13·8 (11·0 - 17·0) | 189 | 57·3 (51·7 - 62·7) | 42 | 12·7 (9·3 - 16·8) | 99 | 30·0 (25·1 - 35·3) |
| | **IPV2** | Pandemic (Timiri n=1099; Jawadhu **n=737)** | 888 | 80·8 (78·3 - 83·1) | 138 | 12·6 (10·7 - 14·7) | 73 | 6·6 (6·2 - 8·3) | 301 | 40·8 (37·3 - 44·5) | 208 | 28·2 (25·0 - 31·6) | 228 | 30·9 (27·6 - 34·4) |
| | | Pre-Pandemic (Timiri n=564; Jawadhu **n=346)** | 442 | 78·4 (74·7 - 81·7) | 84 | 14·9 (12·1 - 18·1) | 38 | 6·7 (4·8 - 9·1) | 184 | 53·2 (47·8 - 58·5) | 82 | 23·7 (19·3 - 28·5) | 80 | 23·1 (18·8 - 27·9) |
| | **Penta3** | Pandemic (Timiri n=1129; Jawadhu **n=756)** | 1001 | 88·7 (86·7 - 90·5) | 80 | 7·1 (5·7 - 8·7) | 48 | 4·3 (3·2 - 5·6) | 414 | 54·8 (51·1 - 58·4) | 117 | 15·5 (13·0 - 18·3) | 225 | 29·8 (26·5 - 33·2) |
| | | Pre-Pandemic (Timiri n=534); Jawadhu **n=327)** | 479 | 89·7 (86·8 - 92·1) | 35 | 6·6 (4·6 - 9·0) | 20 | 3·8 (2·3 - 5·7) | 208 | 63·6 (58·1 - 68·8) | 47 | 14·4 (10·8 - 18·7) | 72 | 22·0 (17·6 - 26·9) |
| **9 months** | **MCV1** | Pandemic (Timiri n=1287)**;** Jawadhu **n=841)** | 451 | 35·0 (32·4 - 37·7) | 648 | 50·4 (47·6 - 53·1) | 188 | 14·6 (12·7 - 16·7) | 74 | 8·8 (6.9 - 10·9) | 403 | 47·9 (44·5 - 51·4) | 364 | 43.3 (39·9 - 46·7) |
| | | Pre-Pandemic Timiri (n=225); Jawadhu n=181) | 51 | 22·7 (17·4 - 28·7) | 158 | 70·2 (63·8 - 76·1) | 16 | 7·1 (4·1 - 11·3) | 17 | 9·3 (5·6 - 14·6) | 118 | 65·2 (57·8 - 72·1) | 46 | 25·4 (9·2 - 32·4) |

**Supplementary Table 5**: Overall delay in vaccination at 6th,10th and 14th week doses in Jawadhu Hills and Timiri during the study period

| | | Jawadhu Hills | | | Timiri | | |
|---|---|---|---|---|---|---|---|
| **Time point** | **Vaccine** | **N** | **n** | **Delayed vaccination (%)** | **N** | **n** | **Delayed vaccination (%)** |
| **6weeks** | OPV1 | 993 | 240 | 24·2 | 1647 | 93 | 5·7 |
| | Rota1 | 918 | 228 | 24·8 | 1549 | 84 | 5·4 |
| | IPV1 | 959 | 228 | 23·8 | 1598 | 93 | 5·8 |
| | Penta1 | 995 | 241 | 24·2 | 1644 | 91 | 5·5 |
| **10 weeks** | OPV2 | 905 | 195 | 21·6 | 1627 | 80 | 4·9 |
| | Rota2 | 835 | 183 | 21·9 | 1531 | 79 | 5·2 |
| | Penta2 | 904 | 192 | 21·2 | 1626 | 78 | 4·8 |
| **14 weeks** | OPV3 | 785 | 160 | 20·4 | 1579 | 116 | 7·4 |
| | Rota3 | 717 | 156 | 21·8 | 1510 | 113 | 7·5 |
| | IPV2 | 775 | 290 | 37·4 | 1552 | 222 | 14·3 |
| | Penta3 | 786 | 164 | 20·9 | 1595 | 115 | 7·2 |
| **9 months** | MCV1 | 612 | 521 | 85·1 | 1308 | 806 | 61·6 |

Supp_Figure_1

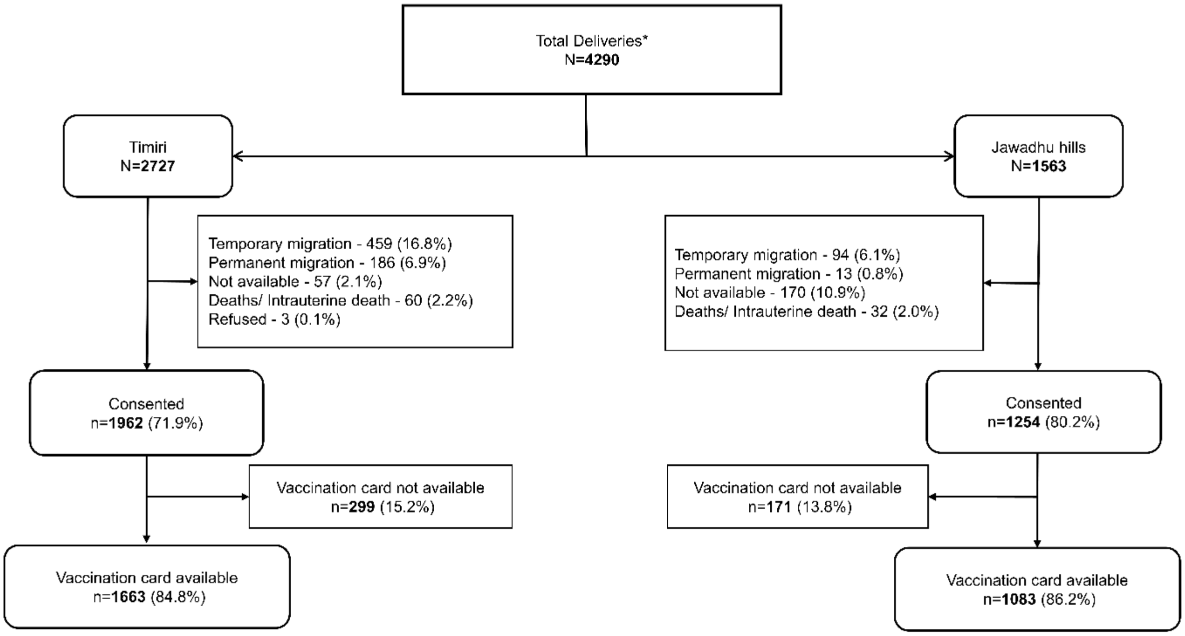

Supp_Figure_2A_2B

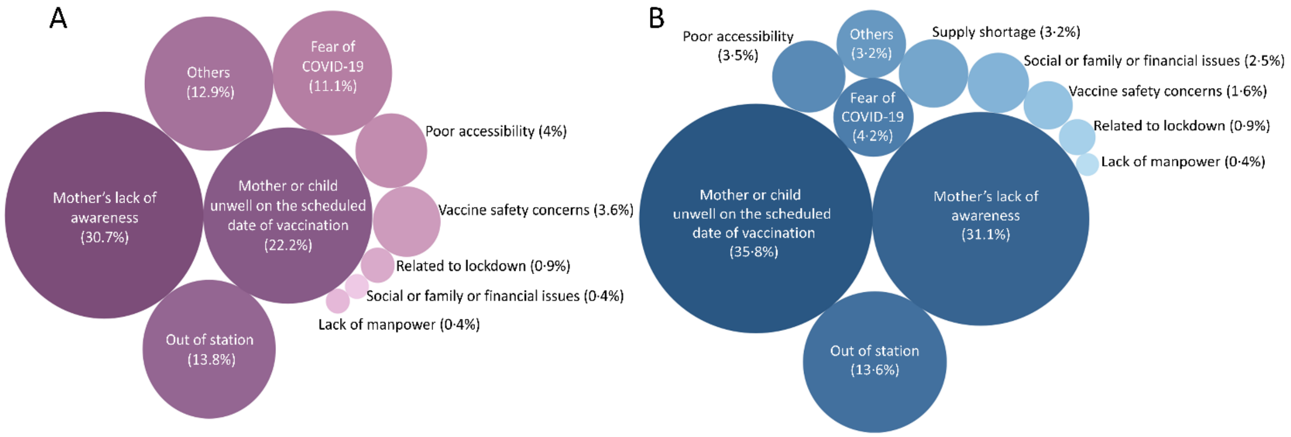
